# Sociodemographic and regional differences in neonatal and infant mortality in Switzerland: The Swiss National Cohort

**DOI:** 10.1101/2023.09.19.23295765

**Authors:** Veronika W Skrivankova, Leonie D Schreck, Claudia Berlin, Radoslaw Panczak, Kaspar Staub, Marcel Zwahlen, Sven M Schulzke, Matthias Egger, Claudia E Kuehni

**Author notes:** **Address for correspondence including e-mail address** Prof. Dr. med. Claudia E. Kuehni Institute of Social and Preventive Medicine Mittelstrasse 43 CH-3012 Bern.

## Abstract

**Background:** Despite a well-funded healthcare system with universal insurance coverage, Switzerland has one of the highest neonatal and infant mortality rates among high-income countries. Identifying avoidable risk factors targeted by evidence-based policies is a public health priority. We describe neonatal and infant mortality in Switzerland from 2011–2018 and explore associations with neonatal and pregnancy-related variables, parental sociodemographic information, regional factors, and socioeconomic position (SEP) using data from a long-term nation-wide cohort study.

**Methods:** We included 680,077 live births—representing 99.3% of all infants born in Switzerland between January 2011 and December 2018. We deterministically linked the national live birth register with the mortality register and with census and survey data to create a longitudinal dataset of neonatal and pregnancy-related variables; parental sociodemographic information, such as civil status, age, religion, education, nationality; regional factors, such as urbanity, language region; and the Swiss neighbourhood index of SEP (Swiss-SEP index). Information on maternal education was available for a random subset of 242,949 infants. We investigated associations with neonatal and infant mortality by fitting multivariable Poisson regression models with robust standard errors. Several sensitivity analyses assessed the robustness of our findings.

**Results:** Overall, neonatal mortality rates between 2011 and 2018 were 3.0 per 1000 live births, varying regionally from 3.2 in German-speaking to 2.4 in French-speaking and 2.1 in Italian-speaking Switzerland. For infant mortality, respective rates were 3.7 per 1000 live births overall, varying from 3.9 to 3.3 and 2.9. Adjusting for sex, maternal age, multiple birth and birth rank, neonatal mortality remained significantly associated with language region [rate ratio (RR) 0.72, 95% confidence interval (CI): 0.64–0.80 for French-speaking and RR 0.66, 95% CI: 0.51–0.87 for Italian-speaking region], with marital status (RR 1.55, 95% CI: 1.40–1.71 for unmarried), nationality (RR 1.40, 95% CI: 1.21–1.62 for non-European Economic Area vs. Swiss), and the Swiss-SEP index (RR 1.17, 95% CI: 1.00–1.36 for lowest vs. highest SEP quintile). In the subset, we showed a possible association of neonatal mortality with maternal education (RR 1.24, 95% CI: 0.95–1.61 for compulsory vs tertiary education).

**Conclusion:** We provide detailed evidence about the social patterning of neonatal and infant mortality in Switzerland and reveal important regional differences with about 30% lower risks in French-and Italian-speaking compared with German-speaking regions. Underlying causes for such regional differences, such as cultural, lifestyle, or healthcare-related factors, warrant further exploration to inform and provide an evidence base for public health policies.

## Manuscript

## Introduction

Switzerland has comparatively high neonatal and infant mortality rates. The Global Burden of Disease Study reported for 2019 a neonatal mortality rate of 2.57 per 1000 live births for Switzerland [1]. Among high-income countries, only the United States (US), Greenland, Canada, and some southern Latin American countries rated worse. In Western Europe, average neonatal mortality was 2.00 per 1000 live births. While the United Kingdom (UK) and Malta were roughly comparable to Switzerland, all neighbouring countries (Austria, France, Germany, and Italy) reported rates around or below 2.00 per 1000 live births. Since Switzerland has a high overall standard of living, universal healthcare with a compulsory health insurance, high government expenditures on health [2], and ranks well with respect to other health indicators such as life expectancy [3], the rates are surprising.

*Social and regional inequalities in health*—health variations not explained by genetic or constitutional factors—highlight the extent of avoidable ill health in a society and show possibilities for better healthcare with more equitable distribution of resources. We previously reported significant spatial variations of birth weight and gestational age in Switzerland, which language region mainly explained, along with additional urbanization, parental nationality, civil status, and altitude [4]. Other studies from Switzerland—focused on parental income and migrant status—showed increased mortality among infants of parents with low incomes, yet reduced risks if mothers originated from European Union (EU) or European Free Trade Association (EFTA) countries compared with Swiss-born mothers [5, 6]. Until now, other social and regional determinants of infant mortality remained uninvestigated in Switzerland.

We describe neonatal and infant mortality for infants born in Switzerland between 2011 and 2018 and examine associations of mortality rates with parental sociodemographic information, regional factors, and socioeconomic position.

## Materials and methods

We used data from the longitudinal dataset of the Swiss National Cohort (SNC) which consists of information from the live birth and mortality register and from census and structural surveys which had been deterministically linked using encrypted Swiss social security numbers as unique national identifiers [7, 8].

### Study population and outcomes

Within the SNC dataset, our study used information on all live births from 1 January 2011–31 December 2018 and followed up until either death, emigration from Switzerland, or survival of 1 year, whichever occurred first. We defined *neonatal death* as death within 28 days and *infant death* as any death within 365 days after live birth. We defined *neonatal mortality rate* (NMR) as the proportion of all neonatal deaths to all live births and *infant mortality rate* (IMR) as the proportion of all infant deaths to all live births.

### Exposures of interest

We first identified variables previously established as important predictors of early mortality or as potential confounders between socioeconomic position (SEP) and early mortality [9–13]. We included neonatal and pregnancy-related variables, such as newborn sex, multiple birth (singletons, twins, etc.), birth rank (sibling order from the same mother), gestational age, and birth weight. Parental sociodemographic factors included parental age, nationality, and education and maternal civil status, place of residence and religion. Regional factors included urbanization level and language region. We recorded SEP using the Swiss neighbourhood index of socioeconomic position (Swiss-SEP index)—an area-based socioeconomic measure of maternal residence [14]. Supplementary **Text S1** describes the Swiss-SEP index, data sources, and variable definitions.

### Statistical analysis

We calculated crude neonatal and infant mortality rates—among all eligible live births and across levels of variables—including Swiss-SEP index quintiles. To better understand relationships between SEP indicators, we plotted binary relationships between the SEP index and other predictors. To assess the effect of sociodemographic and other determinants on mortality, we fitted Poisson regression models [15] with robust standard errors.

We excluded observations with missing values for any used predictors from analyses. For unmarried parents in Switzerland, fathers often register long after birth; therefore, for children who died early, paternal information was more likely missing [missing not at random (MNAR)]; thus, we excluded it from our analyses. Further, we did not adjust for gestational age and birth weight since they possibly reside on causal pathways between SEP and child mortality. Adjusting reduces effects of SEP and may introduce collider-stratification bias [16–18]. We included all remaining predictors in our main model, excepting maternal education and religion. In a second model, we included maternal education; in a third model, maternal religion. Both models were fitted to smaller subsets with available information. All variables were categorical except for maternal age, which we modelled by quadratic spline with a knot at age 30. We calculated credible intervals for RR at selected ages using a bootstrap method with 100 samples. We performed all analyses in statistical software R, version 4.1.1 [19].

We conducted two sensitivity analyses to test the robustness of the main model, one including only singleton births and one adding a sixth SEP category for missing values to compare risk of death in the otherwise excluded group. Observations with missing values for maternal nationality remained excluded (**Text S1**).

## Results

### Characteristics of study population

We linked all deaths from the death register with corresponding live births in the live birth register. From January 2011–December 2018 among 684,716 live births registered in Switzerland, 352,003 (51.4%) were male; 24,440 (3.6%) twins; 682 (0.1%) triplets; and 23 (0.03‰) quadruplets (**Table 1**). Most infant deaths occurred during the first week of life (**Figure 1**). Mean maternal age at birth was 31.4 (standard deviation = 6.2) years. Most mothers (77.2%) were married or in registered partnerships; 959 (0.1%) younger than age 18 at delivery; and 2,385 (0.4%) age 45 or older. Most mothers (60.3%) were Swiss nationals with one-third (29.7%) from other European (7.8% Southern Europe; 12.0% EEA; and 9.9% non-EEA countries), and one-tenth (9.9%) from non-European countries.

**Figure 1:**
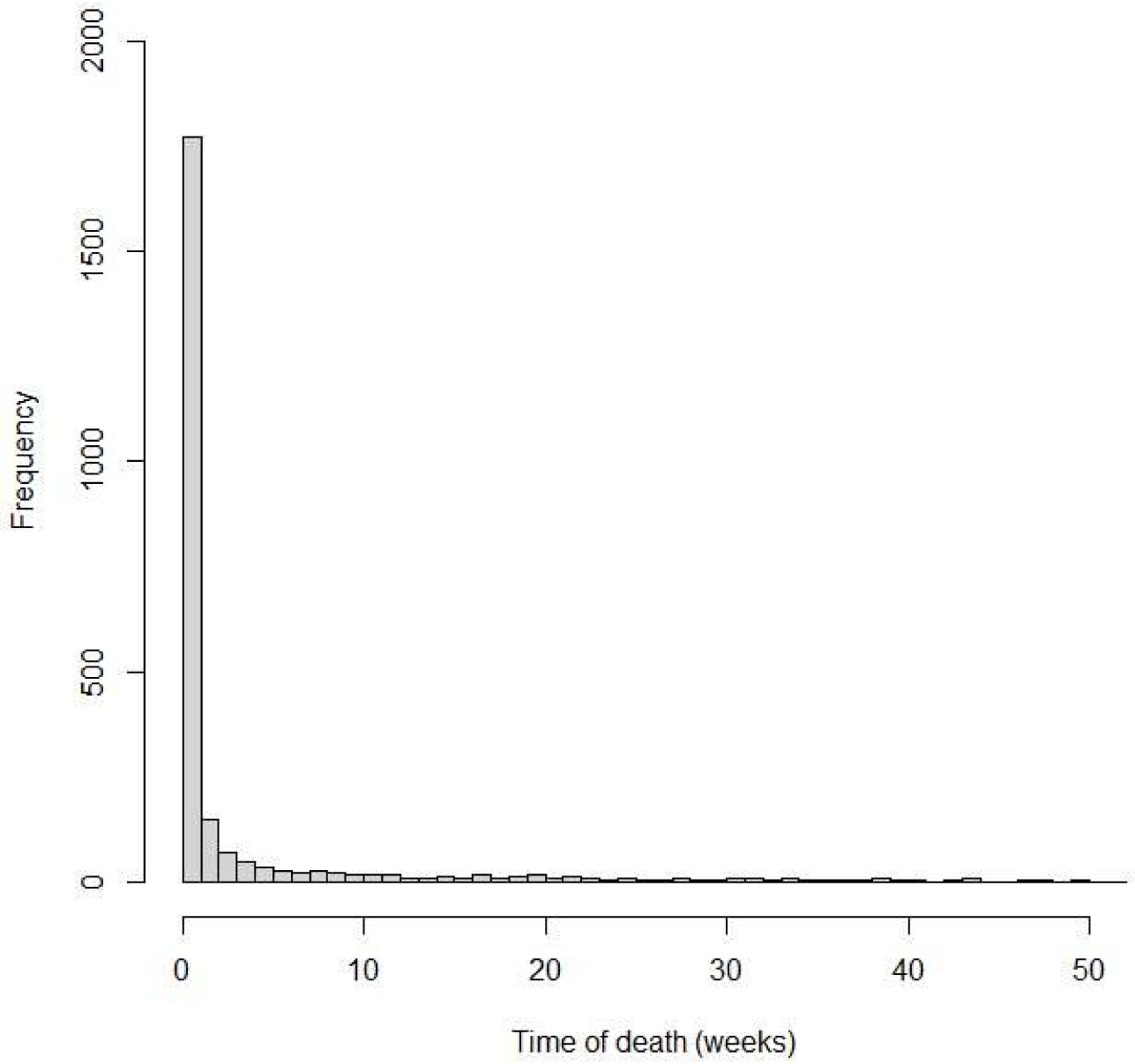
Distribution of time to death among all live births from 2011–2018 registered in Switzerland.

**Table 1.**
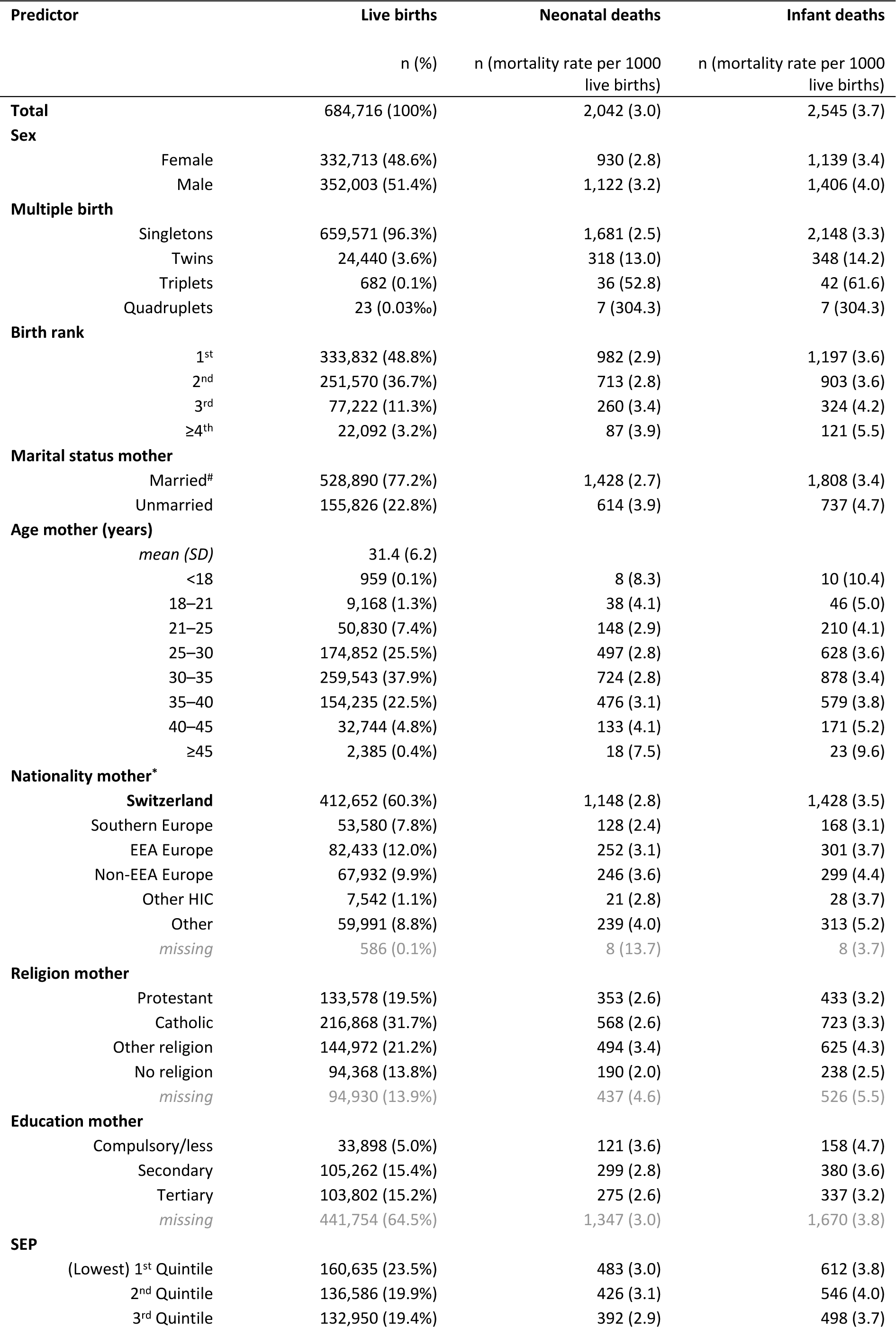
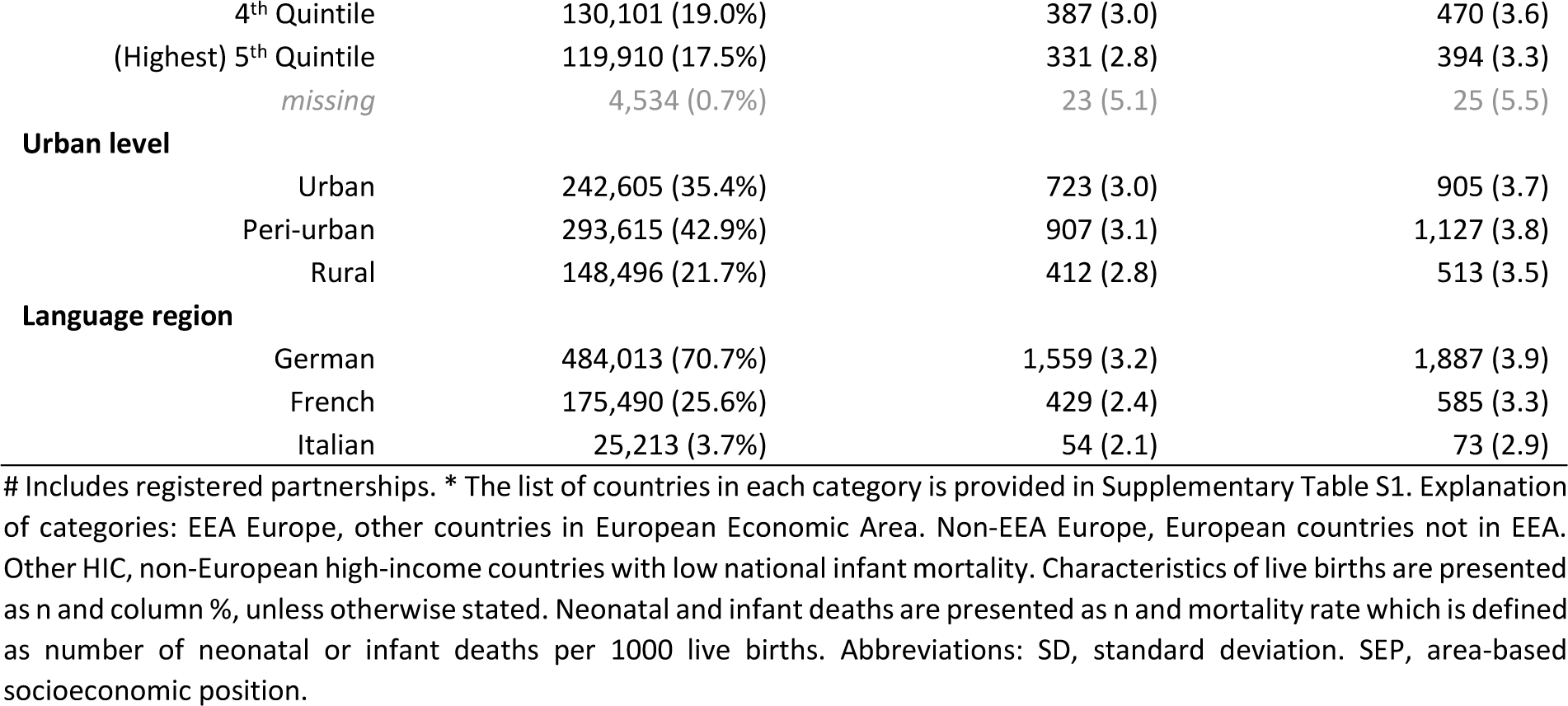
Population characteristics for all live births, number of neonatal and infant deaths, and respective mortality rates in Switzerland in years 2011–2018.

The lowest SEP quintile was slightly overrepresented (23.5%); the highest quintile underrepresented (17.5%). Swiss-SEP index was associated with the proportion of multiple births (A), maternal education (C), maternal nationality (D), and language region (E), and inversely associated with proportion of teenage pregnancies (B) yet not marital status (F) (**Figure 2**).

**Figure 2.**
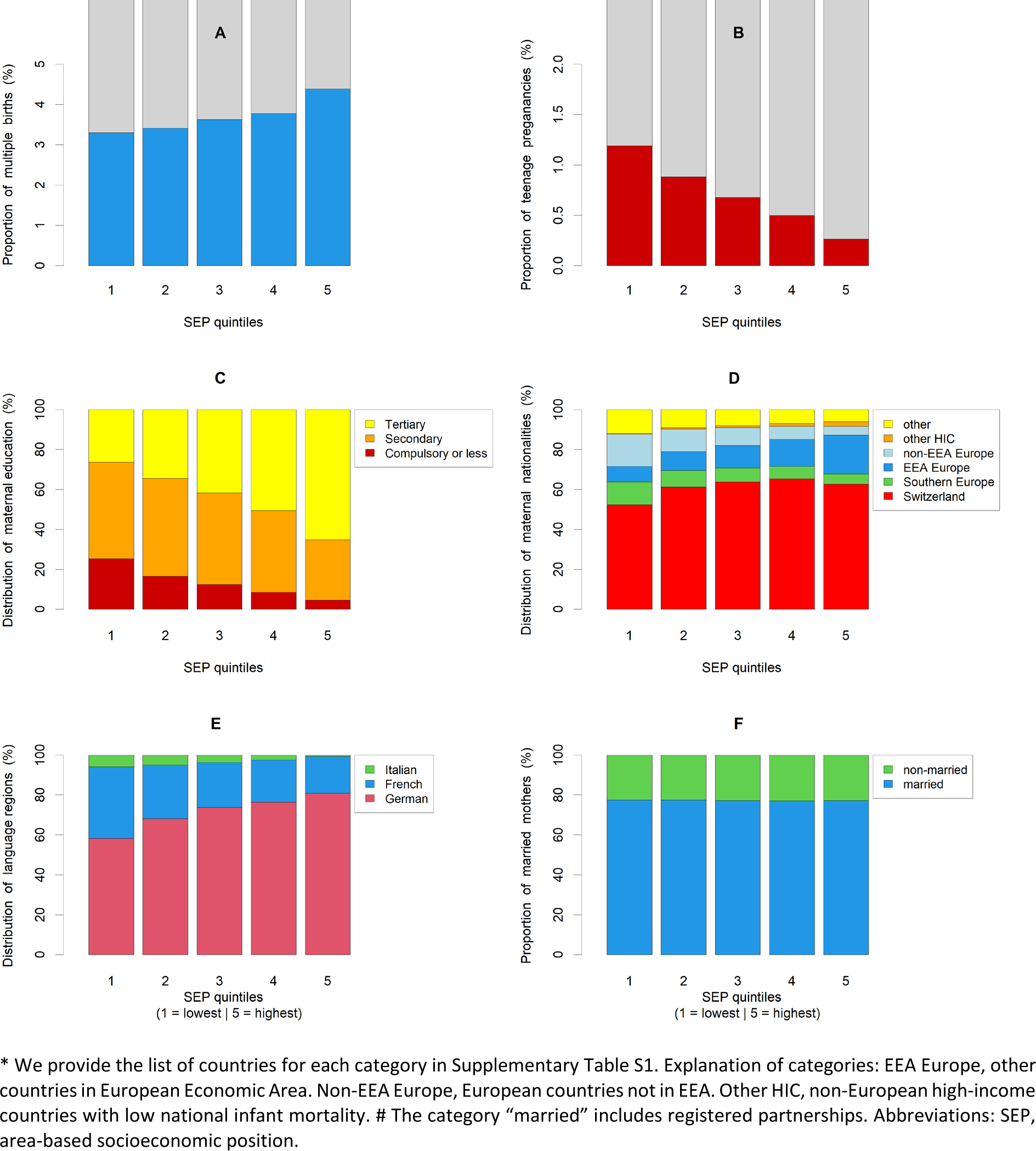
Distribution of selected predictors across SEP quintiles; A) multiple births, B) teenage pregnancies, C) maternal education, D) maternal nationality*, E) language region, and F) marital status^#^.

### Neonatal and infant mortality rate

Among 684,716 live births, 2,042 neonatal deaths occurred within 28 days and 2,545 infant deaths 365 days from birth—an overall NMR of 3.0 and IMR of 3.7 per 1000 live births (**Table 1**). As expected, neonatal and infant mortality rates were higher for males, multiple births, increasing birth rank, and older mothers. Mortality was higher among infants of unmarried mothers (NMR 3.9 per 1000 live births vs 2.7 of married mothers) and varied by nationality—highest for infants of mothers who immigrated from low-income countries outside Europe (NMR 4.0 per 1000 live births vs 2.8 from Swiss mothers). NMR varied regionally from 3.2 in German-speaking to 2.4 in French-speaking and 2.1 in Italian-speaking Switzerland. For infant mortality, respective regional rates varied from 3.9 to 3.3 and 2.9.

We describe unincluded variables in our analyses in supplementary **Table S2**. As expected, gestational age and birth weight were strongly associated with mortality. Differences across paternal age, nationality, and education showed similar patterns—yet, less pronounced—as corresponding maternal characteristics. Mortality rates in live births with missing paternal age and nationality information were 10-times higher than in those with non-missing paternal information—a massive violation of the missing completely at random or MCAR assumption supporting our decision to exclude paternal information from our analyses.

### Predictors of neonatal and infant mortality

Our main multivariable Poisson regression (**Table 2**, **Figure 3**) used data from 680,077 live births, excluding 4,534 (0.7%) with unavailable Swiss-SEP index and 586 with missing maternal nationality (**Figure 4)**. Our results confirm well-known associations with sex, multiple pregnancies, and maternal age, showing a typical U-shaped association (**Figure S1**).

**Figure 3.**
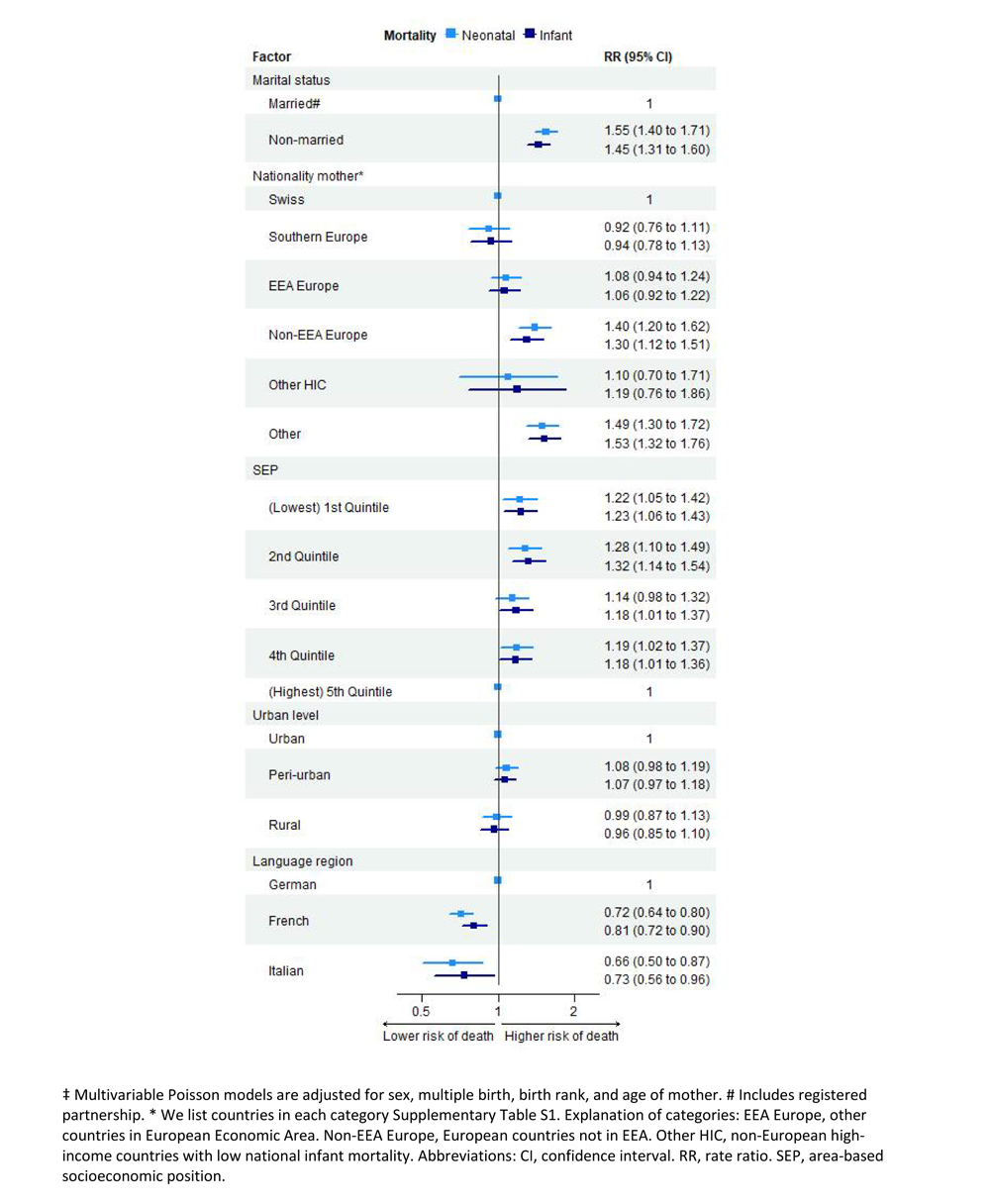
Main analysis: Neonatal and infant mortality rate ratios (RR) in Switzerland 2011–2018, for all live births with available information on all predictors (N = 680,077), based on multivariable Poisson models^‡^.

**Figure 4.**
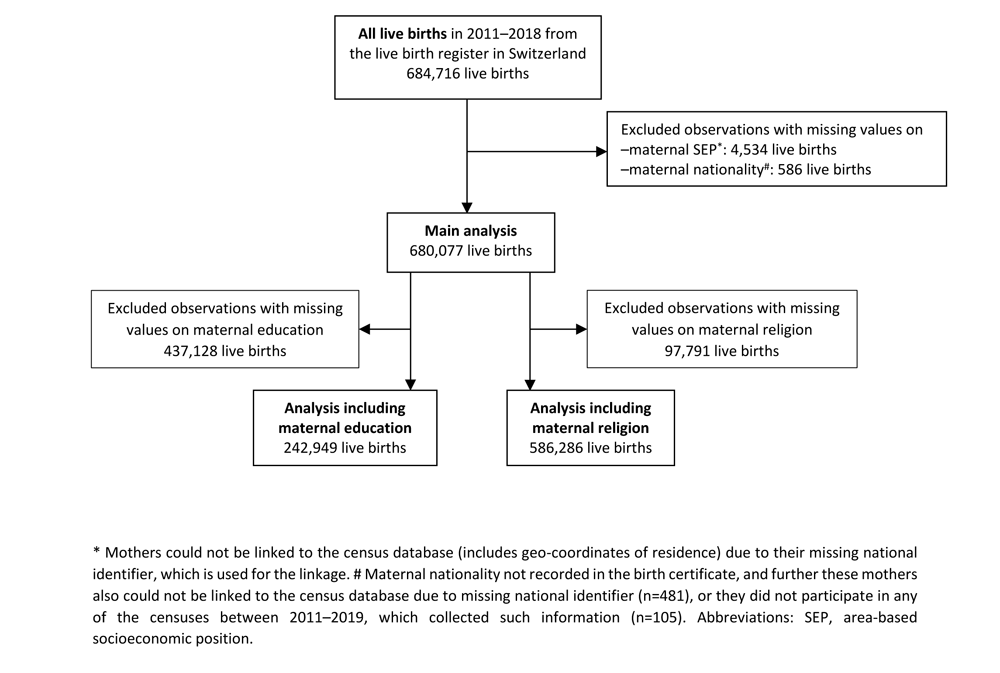
Population flow chart.

**Table 2.** Main analysis: Neonatal and infant mortality rate ratios (RR) in Switzerland 2011–2018, for all live births with available information on all predictors (N = 680,077) based on multivariable Poisson models.

| Predictor | Neonatal mortality |  | Infant mortality |  |
| --- | --- | --- | --- | --- |
|  | RR (95% CI) | p-value | RR (95% CI) | p-value |
| **Sex** |  | 0.003** |  | <0.001*** |
| Female | 1 |  | 1 |  |
| Male | 1.14 (1.05, 1.25) |  | 1.18 (1.09, 1.27) |  |
| **Multiple birth** |  | <0.001*** |  | <0.001*** |
| Singletons | 1 |  | 1 |  |
| Twins | 5.19 (4.58, 5.88) |  | 4.38 (3.89, 4.92) |  |
| Triplets | 20.2 (14.5, 28.2) |  | 17.7 (13.1, 24.1) |  |
| Quadruplets | 63.9 (21.4, 191) |  | 46.7 (15.5, 140) |  |
| **Birth rank** |  | 0.53 |  | 0.39 |
| 1 | 1 |  | 1 |  |
| 2 | 0.93 (0.84, 1.02) |  | 0.98 (0.90, 1.08) |  |
| 3 | 0.98 (0.84, 1.13) |  | 1.04 (0.91, 1.19) |  |
| ≥4 | 0.96 (0.76, 1.22) |  | 1.16 (0.95, 1.43) |  |
| **Marital status** |  | <0.001*** |  | <0.001*** |
| Married# | 1 |  | 1 |  |
| Unmarried | 1.55 (1.40, 1.71) |  | 1.45 (1.32, 1.58) |  |
| **Age of mother<sup>‡</sup>** |  | <0.001*** |  | <0.001*** |
| 15 | 1.85 (1.13, 3.21) |  | 2.09 (1.36, 3.31) |  |
| 18 | 1.42 (1.06, 1.94) |  | 1.58 (1.20, 2.06) |  |
| 20 | 1.25 (1.03, 1.51) |  | 1.36 (1.13, 1.60) |  |
| 25 | 1.01 (0.95, 1.07) |  | 1.06 (1.00, 1.12) |  |
| 30 | 1 |  | 1 |  |
| 35 | 1.12 (1.06, 1.21) |  | 1.08 (1.01, 1.15) |  |
| 40 | 1.29 (1.17, 1.45) |  | 1.26 (1.12, 1.41) |  |
| 45 | 1.55 (1.26, 1.85) |  | 1.59 (1.33, 1.86) |  |
| 50 | 1.89 (1.25, 2.58) |  | 2.20 (1.58, 2.77) |  |
| **Nationality mother\*** |  | <0.001*** |  | <0.001*** |
| Switzerland | 1 |  | 1 |  |
| Southern Europe | 0.92 (0.77, 1.11) |  | 0.94 (0.80, 1.11) |  |
| EEA Europe | 1.07 (0.94, 1.23) |  | 1.06 (0.93, 1.20) |  |
| Non-EEA Europe | 1.40 (1.21, 1.62) |  | 1.30 (1.14, 1.49) |  |
| Other HIC | 1.09 (0.70, 1.71) |  | 1.19 (0.81, 1.75) |  |
| Other | 1.50 (1.30, 1.73) |  | 1.53 (1.35, 1.73) |  |
| **SEP index** |  | 0.06 |  | 0.003** |
| (Lowest) 1<sup>st</sup> quintile | 1.17 (1.00, 1.36) |  | 1.22 (1.06, 1.40) |  |
| 2<sup>nd</sup> quintile | 1.25 (1.08, 1.45) |  | 1.30 (1.14, 1.49) |  |
| 3<sup>rd</sup> quintile | 1.16 (1.00, 1.35) |  | 1.21 (1.05, 1.38) |  |
| 4<sup>th</sup> quintile | 1.11 (0.96, 1.29) |  | 1.12 (0.98, 1.28) |  |
| (Highest) 5<sup>th</sup> quintile | 1 |  | 1 |  |
| **Urban level** |  | 0.21 |  | 0.13 |
| Urban | 1 |  | 1 |  |
| Peri-urban | 1.08 (0.98, 1.19) |  | 1.07 (0.98, 1.17) |  |
| Rural | 0.99 (0.87, 1.13) |  | 0.96 (0.86, 1.08) |  |
| **Language region** |  | <0.001*** |  | <0.001*** |
| German | 1 |  | 1 |  |
| French | 0.72 (0.64, 0.80) |  | 0.81 (0.73, 0.89) |  |
| Italian | 0.66 (0.51, 0.87) |  | 0.73 (0.58, 0.93) |  |

Children of unmarried mothers had higher mortality than those of married mothers (neonatal RR 1.55, 95% CI: 1.40–1.71; infant RR 1.45, 95% CI: 1.32–1.58). Newborns of mothers from non-EEA countries had higher mortality than offspring of Swiss mothers (neonatal RR 1.40, 95% CI: 1.21–1.62; infant RR 1.30, 95% CI: 1.14–1.49); newborns of mothers from outside Europe—except high-income countries—fared worst (neonatal RR 1.50, 95% CI: 1.30–1.73; infant RR 1.53, 95% CI: 1.35–1.73).

Swiss-SEP index was also associated with neonatal and infant mortality, showing approximately 20% higher risks in all SEP quantiles compared with the highest quintile and no evidence for a dose-response relationship. Adjusting for all other factors, mortality rates were lower in French-speaking (neonatal RR 0.72, 95%CI: 0.64–0.80; infant RR 0.81, 95% CI: 0.73–0.89) and Italian-speaking (neonatal RR 0.66, 95% CI: 0.51–0.87; infant RR 0.73, 95% CI: 0.58– 0.93) regions when compared with the German-speaking region. Urbanization was not associated. We show results from unadjusted models in **Table S3** for comparison.

In our first of two secondary analyses, which included a subset of 242,949 live births with available maternal education information, we showed a trend for higher mortality among children of mothers with compulsory education (neonatal RR 1.24, 95% CI: 0.95–1.61; infant RR 1.24, 95% CI: 0.99–1.57) when compared with mothers with finished tertiary education (**Figure 4**, **Table 3**). Associations with all other predictors remained consistent with the main model.

**Table 3.**
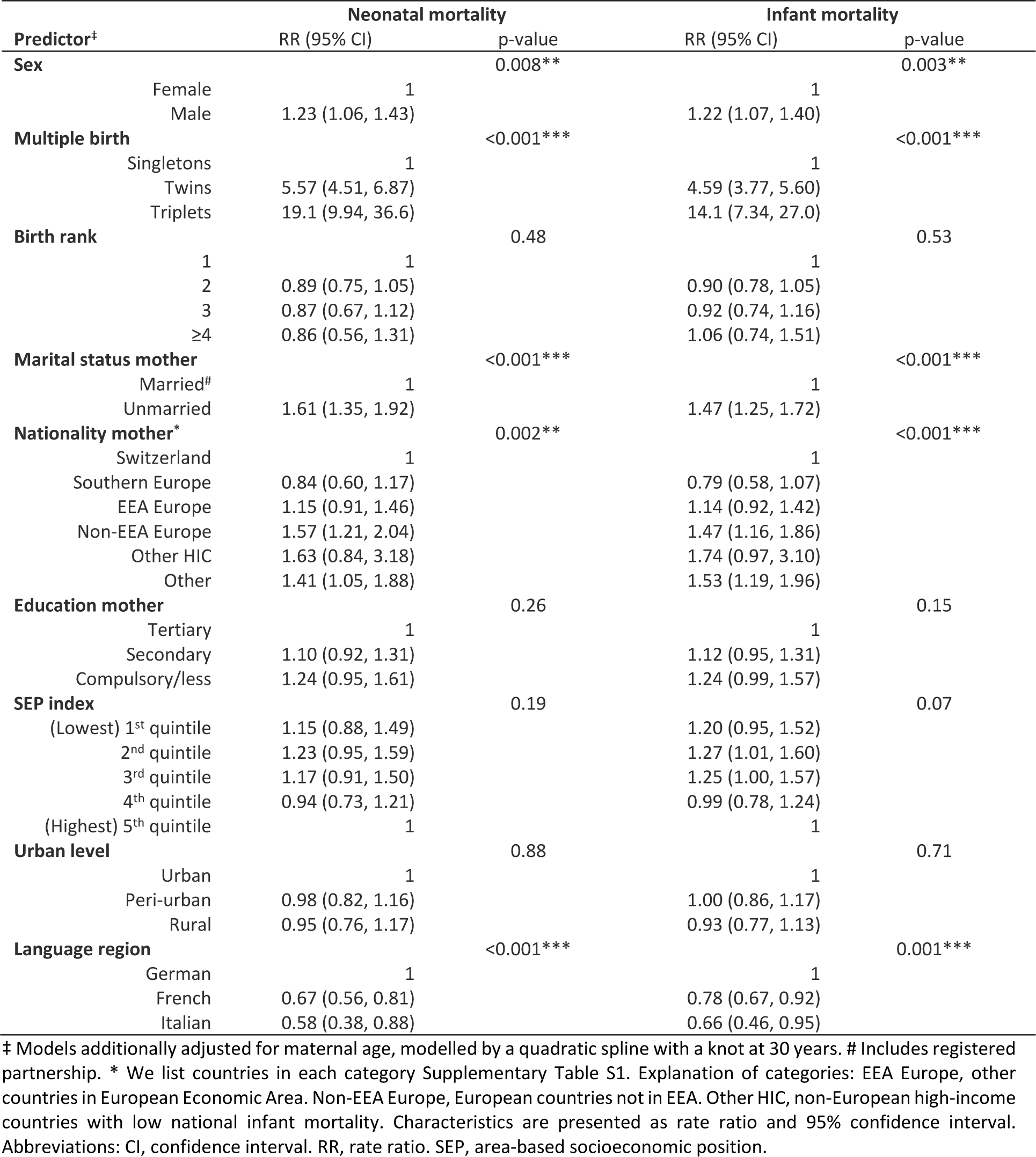
Secondary analysis: Neonatal and infant mortality rate ratios (RR) in Switzerland 2011–2018, for all live births with available information on all predictors including **maternal education** (N=242,949), based on multivariable Poisson models.

Another secondary analysis included a subset of 586,286 live births with available maternal religion information (**Figure 4, Table S4**). We found a strong association between religion and mortality (other religion vs Protestant: neonatal RR 1.31, 95% CI: 1.11–1.55; no religion vs Protestant: neonatal RR 0.76, 95% CI: 0.63–0.91), yet no difference between Protestant and Catholic denomination (Catholic vs Protestant: neonatal RR 1.05, 95% CI: 0.91– 1.20). Associations with other predictors, including language region, remained largely consistent with the main model. We show population characteristics of subsets and characteristics of live births we excluded from our main analyses in the supplementary material (**Table S5**). Results from sensitivity analyses remained similar to our main results (**Tables S6 and S7**).

## Discussion

### Summary of findings

Based on a longitudinal dataset of all children born alive in Switzerland between 2011 and 2018, we confirmed neonatal mortality (3.0 per 1000 live births) and infant mortality (3.7 per 1000 live births) is higher in Switzerland than most other high-income countries. Adjusting for well-described associations with sex, multiple births, and maternal age, we found additional variation by sociodemographic and socioeconomic factors, with higher mortality for mothers who are unmarried, immigrated from low-income countries, and living outside the top quintile of the Swiss-SEP index. Even after adjusting for these factors, a strong association with language region—lower neonatal and infant mortality among inhabitants of French-and Italian-speaking regions—remained.

### Strengths and weaknesses

Ours is the largest of few studies on neonatal outcomes in Switzerland—it also includes the most comprehensive socioeconomic information. We linked the dataset deterministically, resulting in a cohort study with few overall missing data. Our considerable sample size enabled multivariable analysis and allowed modelling associations, such as maternal age, carefully. We did deliberately not adjust our regression analyses for intermediates, such as birth weight and gestational age, which may be important drivers of associations of interest. Adjustment would have artificially reduced the effect of SEP or worse, induced collider bias—a mechanism described repeatedly, using examples such as “birth weight paradox” [20–23].

Our study also displays weaknesses. Although we used information from eight consecutive years, the limited dataset size prevented multivariable analysis of postneonatal deaths (n = 503). Further, paternal data were missing for unmarried mothers when children died early. We thus refrained from modelling associations with paternal factors. However, most previous studies showed stronger effects of maternal compared with paternal SEP [24]. For mothers, we included comprehensive sociodemographic, regional, and socioeconomic data, yet lacked professional and environmental exposure (e.g., air pollution, volatile other organic compounds) information on lifestyle; health behaviours, such as smoking and alcohol consumption; and maternal health, such as obesity, gestational diabetes, preeclampsia, or hypertension. Without professional and environmental exposure information, we faced limited possibilities to explore causal pathways. We also lacked information about healthcare visits, prenatal screening, and post-natal care. Finally, an important limitation our study shares with others relates to the so-called “live-birth bias” [23, 25]. We lacked stillbirth and late pregnancy termination data for severe foetal conditions. Although harmonized definitions of “late pregnancy termination,” “stillbirth,” and “live birth” followed by “early neonatal death” for Swiss vital statistics are the same across Switzerland, there might be regional differences in how these distinctions are handled in real life.

### How do results compare to other studies

Few studies investigated social determinants of neonatal and infant mortality in Switzerland. The Federal Statistical Office publishes routine data from single registers, yet datasets remain unlinked and multivariable analyses unperformed [26, 27]. Previous research focused on migrant populations and country origins. For example, a vital statistics analysis reported for 1980–2011, 29% higher NMR and up to 18% higher IMR among children with non-Swiss nationalities [6]. A follow-up study for 2011–2017 linked live births, infant deaths, and parental income from the Swiss Central Compensation Office [5] and found increased risk for infants of mothers with low-income. Associations with maternal country of birth were less clearcut: IMR was lower for mothers born in EU or EFTA (OR 0.83, 95% CI: 0.71–0.97) and higher (OR 1.15, 95% CI: 1.01–1.30) if born in non-OECD countries. IMR among asylum seekers was unexpectedly low (OR 0.57, 95% CI: 0.36–0.91) compared with residence permit holders.

More information is available from other countries, particularly the UK—a country also with a high NMR and IMR [28–31]. A systematic review of 35 UK-based studies reported increased odds of stillbirth, perinatal and neonatal mortality, preterm birth, and low birth weight for women from lower levels of social class [28]. Zylberszteijn et al. compared neonatal, postneonatal, and child mortality between England and Sweden; they attributed 77% of excess neonatal mortality in England to birth characteristics (gestational age, birth weight, sex and congenital malformations) and only 3% to socioeconomic factors, which included maternal age [31]. It is likely that this study has underestimated the role of SEP, since some SEP effects are mediated through gestational age and birthweight, which they included in their models. Studies from other countries obtained similar findings. In Michigan (US), SEP and maternal risk behaviours explained nearly one-third infant mortality disparity [32]. In Spain, stillbirth risk was doubled for mothers with secondary or lower education and among mothers from African countries [11, 33]. Studies from The Netherlands [34–37], Italy [10, 38, 39], and Canada [13, 40] also reported consistent findings based on individual patient data or ecological studies.

### What does it mean – implications for policy and further research

Overall, we found some variation by socioeconomic factors: less than we expected. In addition, we found consistent, somewhat surprising differences between regions. Although Switzerland maintains a long tradition of peaceful cohabitation between people speaking German, French, or Italian as mother tongues, cultural, social, and behavioural factors, including diet, tobacco smoking and alcohol consumption, voting behaviour at referendums, social policies, and organization of public healthcare vary by region [4, 41–44]. Interpreting our findings requires nuance and further research.

Several mechanisms possibly explain lower risk of neonatal mortality in French-and Italian-speaking Switzerland. First, maternal and foetal health are possibly better from a truly lower incidence of chromosomal abnormalities, severe congenital malformations, and pregnancy complications. However, we consider it rather unlikely—we found lower rather than higher birth weight and gestational age in French and Italian language regions, endpoints often taken as proxy for general foetal and maternal health [4]. Second, even if regions showed similar underlying maternal and foetal health, regions possibly report stillbirth registrations and early neonatal deaths slightly differently. For example, neonates with anencephaly might be reported as stillbirth or as neonatal death. Third, offers or uptake of prenatal screening and decisions or methods for terminating pregnancy possibly vary. In Switzerland, one-third of neonatal deaths before 28 weeks gestation are due to late pregnancy terminations for medically-approved imminent maternal harm or maternal major psychological distress about foetal problems with poor prognoses, such as congenital malformations and chromosomal aberrations [45]. Under such circumstances, infants are stillborn if feticide is conducted prior to birth, yet if pregnancy termination is solely performed by inducing labour, infants may be born alive and die shortly thereafter [46]. Although live births are registered for all infants with signs of life—independent of gestational age or birth weight—neonatal deaths are only reported from 22 weeks gestation or for birth weights above 500 grams. Given 63% of neonatal deaths in our dataset occurred before 28 weeks gestation (**Table S2**), regional practice variations combined with primary resuscitation differences for preterm infants born at limits of viability perhaps strongly influenced neonatal mortality rates [47].

We thus suggest systematic differences in pregnancy-related policies and actions by parents and physicians possibly explain at least part of our findings. This hypothesis is also supported by associations with religion and lower neonatal mortality among couples reporting “no religion.” We could reasonably hypothesize such parents as more likely to terminate pregnancies with lethal malformations.

We suggest future studies using routine administrative data ideally integrate information about stillbirths and late pregnancy terminations. Larger datasets would allow for differentiating between risk factors for perinatal, late neonatal, and postneonatal death. Strong evidence shows social patterning differs between these two outcome measures, with SEP explaining mainly postneonatal deaths, particularly sudden infant death syndrome [13, 48]. Including more information about birth mechanism, such as Caesarean section or instrumental deliveries, and health behaviours, such as smoking, in routine Swiss statistics, would further improve exploration of causal pathways. Finally, we need qualitative interview studies with healthcare providers, policy makers, and parents in different language regions and within population strata to shed light on potential mechanisms and develop policies.

### Conclusions

Overall, our national cohort study confirmed high neonatal and infant mortality rates in Switzerland, which showed some variation by sociodemographic and socioeconomic factors, such as nationality, civil status, and area-based socioeconomic position, and revealed robust differences between language regions. While infant mortality in German-speaking regions was high (NMR 3.2 per 1000 live births), rates in French-and Italian-speaking regions were more comparable to other high-income countries (NMR 2.4 and 2.1 per 1000 live births). Further elucidation of relevant pathways and mechanisms is needed to develop evidence-based policies for improving maternal and child health in Switzerland.

## Data Availability

Data are available upon reasonable request. Data may be obtained from a third party and are not publicly available.

## Acknowledgments

We thank Kristin Marie Bivens for her editorial work and guidance on our manuscript.

## Ethics approval

The Swiss National Cohort has been approved by the Ethics Committee of the Canton of Bern (Switzerland).

## Supplementary Materials

**Text S1:** Supplementary material and methods

### Description of the Swiss index of socioeconomic position

The Swiss index of socioeconomic position (Swiss-SEP index) is centred on the level of residence buildings and uses information from a neighbourhood’s 50 closest households. Variables used to construct the index are median rent per square meter, the proportion of households headed by a person in a manual or unskilled occupation, the proportion headed by a person with primary education or less, and the mean number of persons per room, obtained from the 2000 annual census and updated for newly constructed buildings using data from the population surveys in 2012–2015 [13].

### Data sources

The live birth register contains information about a newborn infant’s date of birth, sex, multiple birth, birth rank, gestational age, birth weight, maternal civil status, and residential municipality, parental ages, and nationalities at the time of birth. Urbanization level and language region were based on the maternal residential municipality and obtained from national municipality registers (“Raumgliederung”). As these registers might slightly change from year to year, we matched the year of municipality register to the year of birth. Follow-up time was one year (365 days) for most infants unless they emigrated or died. We obtained the emigration data from the national census database and date of death from the death register. We extracted parental education level from annual national structural surveys and also as available for a random subset of infants whose parents were invited to one of the surveys conducted between 2010 and 2019. In case parents participated in more than one of these structural surveys, we used information from the most recent year. We extracted exact maternal place of residence (geographical coordinates assigned to the address) from the national census database. As official geocoding of residential buildings also slightly changes from year to year (old buildings are extended and new buildings are added), we always used the coordinates from the year of birth. Geographical coordinates of place of maternal residence were then matched with closest available geographical points in the Swiss socioeconomic position (SEP) registry to obtain the Swiss-SEP index [15].

### Variable definitions

#### Neonatal and pregnancy-related variables

Multiple births were categorized as singletons, twins, triplets, or quadruplets. Birth rank was categorized as 1st, 2nd, 3rd, and ≥4th.

#### Parental sociodemographic factors

Civil status of mother was dichotomized as “married” or “unmarried,” with registered partnerships included in the “married” category. Nationality of the mother, as in other publications [6], was categorized as “Switzerland,” “Southern Europe,” “EEA Europe” (other countries in European Economic Area), “Non-EEA Europe” (European countries not in EEA), “Other HIC” (non-European high-income countries, with low national infant mortality) and “Other” (supplementary Table S1). Maternal education referred to the level of finished education at the time when the mother participated in a structural survey between 2011-2019, and was categorized into “compulsory or less,” “secondary,” and “tertiary.” For mothers age 15 or younger at the time of birth, we replaced any missing values with level “compulsory or less.” Religion was categorized as “Protestant,” “Catholic” (Roman Catholic), “Other religion” (Christian Catholic, other Christian, Jewish, Islamic, other religion) and “No religion.”

#### Regional factors

Language regions were “German,” “French,” and “Italian.” They were delimited at the level of the municipalities and follow the definition of the language regions by the Federal Statistical Office, which are based on the results of the federal censuses. Romansh-speaking communities were merged with the category “Italian,” as their frequency was very low (0.2%), and they are geographically closest to the Italian-speaking regions. Levels of urbanization were defined as “urban,” “peri-urban” and “rural.”

#### Socioeconomic position

At the time of development of the Swiss-SEP index, authors calculated its percentiles based on the source Swiss population. For our analyses, we aggregated the SEP index into 5 groups based on quintiles of this underlying SEP distribution.

**Table S1:** List of countries in categories of maternal nationality.

| Region | Countries |
| --- | --- |
| **Switzerland** | Switzerland |
| **Southern Europe** | Greece, Italy, Portugal, Spain |
| **EEA Europe** (other countries in European Economic Area) | Austria, Belgium, Bulgaria, Cyprus, Czech Republic, Denmark, Estonia, Finland, France, Germany, Ireland, Iceland, Lichtenstein, Latvia, Lithuania, Luxemburg, Malta, Netherlands, Norway, Poland, Romania, Slovakia, Slovenia, Sweden, United Kingdom* |
| **Non-EEA Europe** (European countries not in EEA) | Albania, Belarus, Bosnia & Herzegovina, Croatia, Hungary, Kosovo, Macedonia, Moldova, Montenegro, Russia, Serbia, Turkey, Ukraine |
| **Other HIC** (non-European high-income countries) | Australia, Canada, Israel, Japan, New Zealand, Singapore, South Korea, Taiwan, United States of America |
| **Other** (other non-European low- and middle-income countries; listed top 12 most represented) | Afghanistan, Eritrea, Brazil, China, India, Iraq, Morocco, Somalia, Sri Lanka, Syria, Thailand, Tunisia, ... |

\* Prior to 2020

**Table S2:** Additional population characteristics for all live births in Switzerland in years 2011–2018.

| Predictor | Live births | Neonatal deaths | Infant deaths |
| --- | --- | --- | --- |
|  | Number (%)<br>or mean (SD) | n (mortality rate per 1000<br>live births) | n (mortality rate per 1000<br>live births) |
| **Total** | 684,716 (100%) | 2,042 (3.0) | 2,545 (3.7) |
| **Gestational age (weeks)** |  |  |  |
| <20<sup>0</sup> | 162 (0.02%) | 162 (1000) | 162 (1000) |
| 20<sup>0</sup>–21<sup>6</sup> | 225 (0.03%) | 222 (986.7) | 223 (991.1) |
| 22<sup>0</sup>–23<sup>6</sup> | 541 (0.08%) | 505 (933.5) | 509 (940.9) |
| 24<sup>0</sup>–25<sup>6</sup> | 774 (0.11%) | 302 (390.2) | 321 (414.7) |
| 26<sup>0</sup>–27<sup>6</sup> | 1,093 (0.16%) | 105 (96.1) | 124 (113.4) |
| 28<sup>0</sup>–29<sup>6</sup> | 1,635 (0.24%) | 67 (41.0) | 82 (50.2) |
| 30<sup>0</sup>–31<sup>6</sup> | 2,692 (0.39%) | 62 (23.0) | 83 (30.8) |
| 32<sup>0</sup>–34<sup>6</sup> | 12,444 (1.82%) | 118 (9.5) | 151 (12.1) |
| 35<sup>0</sup>–36<sup>6</sup> | 29,543 (4.31%) | 96 (3.2) | 139 (4.7) |
| 37<sup>0</sup>–38<sup>6</sup> | 180,756 (26.4%) | 168 (0.9) | 322 (1.8) |
| 39<sup>0</sup>–41<sup>6</sup> | 448,665 (65.5%) | 219 (0.5) | 406 (0.9) |
| ≥42<sup>0</sup> | 3,588 (0.52%) | 4 (1.1) | 6 (1.7) |
| *missing* | 2,598 (0.38%) | 12 (4.6) | 17 (6.5) |
| **Birth weight (grams)** |  |  |  |
| *mean (SD)* | 3290 (549) |  |  |
| <400 | 407 (0.06%) | 373 (916.5) | 373 (916.5) |
| 400–600 | 700 (0.10%) | 520 (742.9) | 540 (771.4) |
| 600–800 | 938 (0.14%) | 302 (322.0) | 323 (344.3) |
| 800–1000 | 1,130 (0.17%) | 105 (92.9) | 119 (105.3) |
| 1000–1200 | 1,105 (0.16%) | 48 (43.4) | 57 (51.6) |
| 1200–1500 | 2,575 (0.38%) | 67 (26.0) | 87 (33.8) |
| 1500–2000 | 8,628 (1.26%) | 101 (11.7) | 132 (15.3) |
| 2000–2500 | 28,933 (4.23%) | 114 (3.9) | 168 (5.8) |
| 2500–3000 | 122,128 (17.8%) | 135 (1.1) | 240 (2.0) |
| 3000–3500 | 272,677 (39.8%) | 146 (0.5) | 286 (1.0) |
| 3500–4000 | 191,153 (27.9%) | 75 (0.4) | 148 (0.8) |
| 4000–4500 | 46,684 (6.82%) | 32 (0.7) | 42 (0.9) |
| ≥4500 | 5,172 (0.76%) | 5 (1.0) | 7 (1.4) |
| *missing* | 2,486 (0.36%) | 19 (7.6) | 23 (9.3) |
| **Age father (years)** |  |  |  |
| *mean (SD)* | 34.5 (6.2) |  |  |
| <18 | 230 (0.03%) | 1 (4.3) | 1 (4.3) |
| 18–21 | 2299 (0.34%) | 2 (0.9) | 5 (2.2) |
| 21–25 | 20406 (2.98%) | 50 (2.5) | 68 (3.3) |
| 25–30 | 111,921 (16.3%) | 276 (2.5) | 362 (3.2) |
| 30–35 | 224,307 (32.8%) | 507 (2.3) | 641 (2.9) |
| 35–40 | 186,592 (27.3%) | 464 (2.5) | 581 (3.1) |
| 40–45 | 84,726 (12.4%) | 235 (2.8) | 297 (3.5) |
| ≥45 | 41,329 (6.04%) | 153 (3.7) | 202 (4.9) |
| *missing* | 12,906 (1.88%) | 354 (27.4) | 388 (30.1) |
| **Nationality father\*** |  |  |  |
| Switzerland | 405,364 (59.2%) | 969 (2.4) | 1,237 (3.1) |
| Southern Europe | 70,341 (10.3%) | 134 (1.9) | 176 (2.5) |
| EEA Europe | 78,115 (11.4%) | 202 (2.6) | 250 (3.2) |
| Non-EEA Europe | 67,309 (9.83%) | 235 (3.5) | 287 (4.3) |
| Other HIC | 5,344 (0.78%) | 8 (1.5) | 13 (2.4) |
| Other | 44,976 (6.57%) | 139 (3.1) | 193 (4.3) |
| *missing* | 13,268 (1.94%) | 355 (26.8) | 389 (29.3) |
| **Education father** |  |  |  |
| Compulsory/less | 30,782 (4.50%) | 95 (3.1) | 121 (3.9) |
| Secondary | 88,815 (13.0%) | 218 (2.5) | 281 (3.2) |
| Tertiary | 120,089 (17.5%) | 297 (2.5) | 386 (3.2) |
| *missing* | 445,030 (65.0%) | 1,432 (3.2) | 1,757 (3.9) |

\* We list countries in each category in Supplementary Table S1. Explanation of categories: EEA Europe, other countries in European Economic Area. Non-EEA Europe, European countries not in EEA. Other HIC, non-European high-income countries with low national infant mortality. Abbreviations: SD, standard deviation.

**Figure S1.**
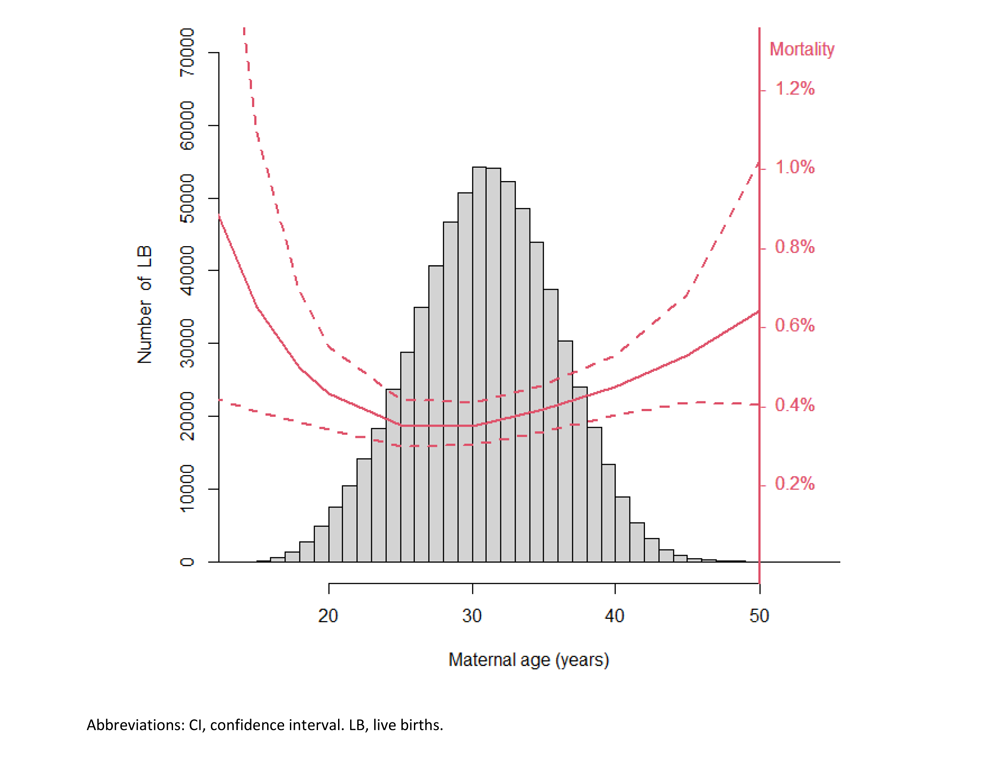
Infant mortality as a function of maternal age at birth. Fitted curve (and the 95% CI) based on the main Poisson model, using a quadratic spline with a knot at 30 years. Mortality rates evaluated at baseline values for all other variables.

**Table S3.** **Univariable**: Neonatal and infant mortality rate ratios (RR) in Switzerland 2011–2018, for all live births with available information on all predictors (N = 680,077), based on **univariable** Poisson models.

| Predictor | Neonatal mortality |  | Infant mortality |  |
| --- | --- | --- | --- | --- |
|  | RR (95% CI) | p-value | RR (95% CI) | p-value |
| **Sex** |  | <0.001*** |  | <0.001*** |
| Female | 1 |  | 1 |  |
| Male | 1.13 (1.04, 1.24) |  | 1.17 (1.08, 1.26) |  |
| **Multiple birth** |  | <0.001*** |  | <0.001*** |
| Singletons | 1 |  | 1 |  |
| Twins | 5.11 (4.53, 5.76) |  | 4.37 (3.90, 4.90) |  |
| Triplets | 20.9 (15.0, 29.1) |  | 19.1 (14.0, 25.9) |  |
| Quadruplets | 61.7 (19.9, 191) |  | 48.2 (15.5, 150) |  |
| **Birth rank** |  | 0.02* |  | <0.001*** |
| 1 | 1 |  | 1 |  |
| 2 | 0.97 (0.88, 1.07) |  | 1.00 (0.92, 1.09) |  |
| 3 | 1.14 (1.00, 1.31) |  | 1.17 (1.03, 1.32) |  |
| ≥4 | 1.29 (1.03, 1.61) |  | 1.47 (1.22, 1.78) |  |
| **Marital status** |  | <0.001*** |  | <0.001*** |
| Married# | 1 |  | 1 |  |
| Unmarried | 1.44 (1.31, 1.58) |  | 1.36 (1.25, 1.49) |  |
| **Age of mother†** |  | <0.001*** |  | <0.001*** |
| 15 | 2.64 (1.34, 4.25) |  | 2.79 (1.45, 4.35) |  |
| 18 | 1.79 (1.18, 2.35) |  | 1.90 (1.27, 2.46) |  |
| 20 | 1.47 (1.11, 1.74) |  | 1.54 (1.19, 1.81) |  |
| 25 | 1.05 (0.99, 1.11) |  | 1.10 (1.05, 1.16) |  |
| 30 | 1 |  | 1 |  |
| 35 | 1.13 (1.06, 1.21) |  | 1.10 (1.04, 1.16) |  |
| 40 | 1.41 (1.26, 1.56) |  | 1.38 (1.27, 1.52) |  |
| 45 | 1.96 (1.57, 2.31) |  | 2.01 (1.72, 2.32) |  |
| 50 | 2.99 (1.96, 4.01) |  | 3.39 (2.43, 4.33) |  |
| **Nationality mother\*** |  | <0.001*** |  | <0.001*** |
| Switzerland | 1 |  | 1 |  |
| Southern Europe | 0.87 (0.73, 1.05) |  | 0.92 (0.78, 1.08) |  |
| EEA Europe | 1.11 (0.97, 1.28) |  | 1.07 (0.95, 1.22) |  |
| Non-EEA Europe | 1.31 (1.14, 1.50) |  | 1.28 (1.13, 1.45) |  |
| Other HIC | 1.02 (0.66, 1.59) |  | 1.11 (0.76, 1.62) |  |
| Other | 1.47 (1.28, 1.69) |  | 1.55 (1.37, 1.75) |  |
| **SEP index** |  | 0.83 |  | 0.28 |
| (Lowest) 1<sup>st</sup> quintile | 1.05 (0.91, 1.20) |  | 1.14 (1.00, 1.29) |  |
| 2<sup>nd</sup> quintile | 1.14 (0.99, 1.32) |  | 1.22 (1.07, 1.39) |  |
| 3<sup>rd</sup> quintile | 1.09 (0.95, 1.26) |  | 1.15 (1.01, 1.32) |  |
| 4<sup>th</sup> quintile | 1.06 (0.92, 1.23) |  | 1.09 (0.95, 1.24) |  |
| (Highest) 5<sup>th</sup> quintile | 1 |  | 1 |  |
| **Urban level** |  | 0.16 |  | 0.12 |
| Urban | 1 |  | 1 |  |
| Peri-urban | 1.05 (0.95, 1.16) |  | 1.04 (0.95, 1.13) |  |
| Rural | 0.94 (0.83, 1.06) |  | 0.93 (0.84, 1.04) |  |
| **Language region** |  | <0.001*** |  | <0.001*** |
| German | 1 |  | 1 |  |
| French | 0.76 (0.68, 0.85) |  | 0.86 (0.78, 0.94) |  |
| Italian | 0.67 (0.51, 0.88) |  | 0.75 (0.59, 0.95) |  |

**Table S4.**
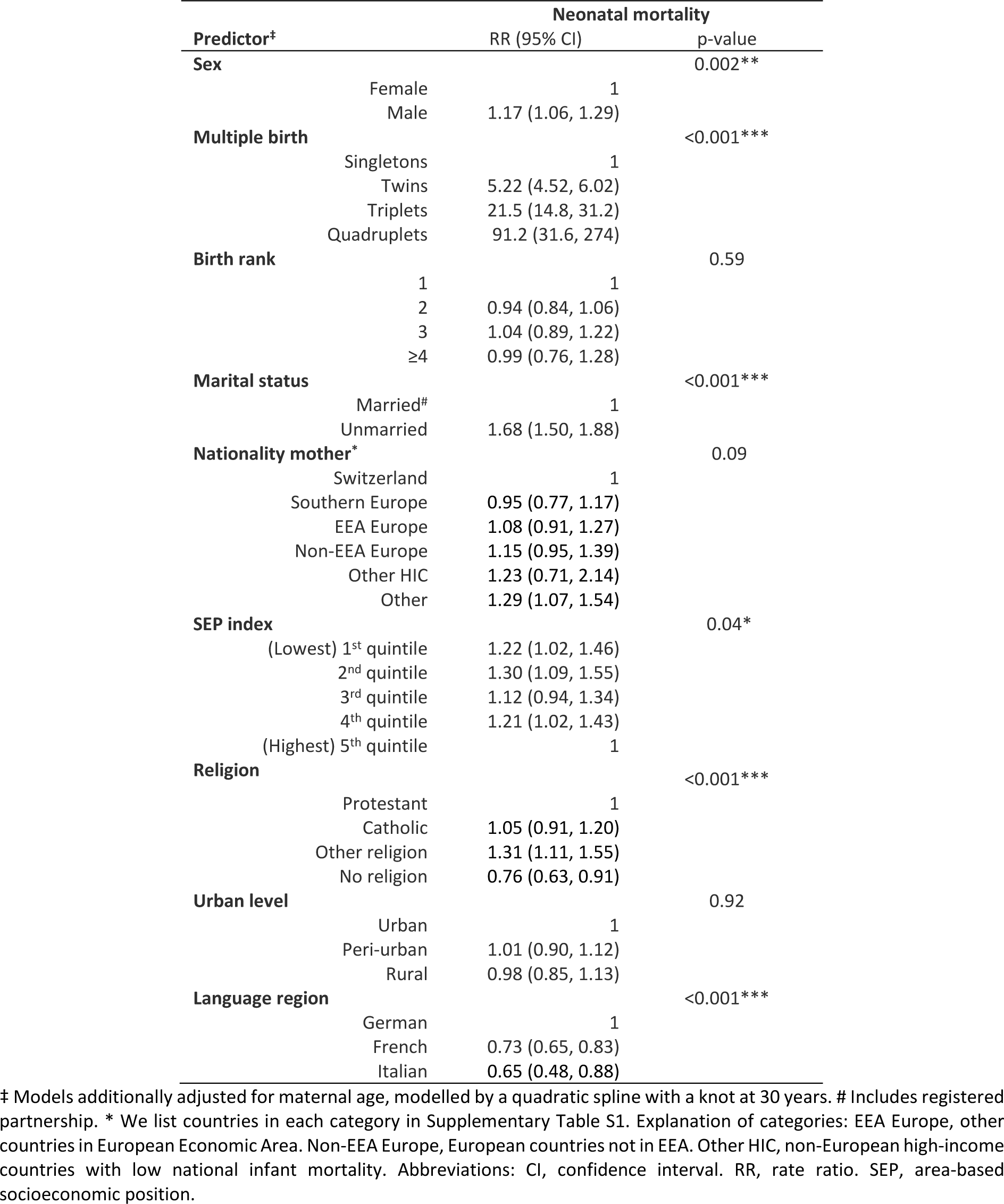
**Secondary analysis**: Neonatal mortality rate ratios (RR) in Switzerland 2011–2018, for all live births with available information on all predictors including **religion** (N = 586,286) based on multivariable Poisson models.

| Predictor<sup>‡</sup> |  | Neonatal mortality |  |
| --- | --- | --- | --- |
|  |  | RR (95% CI) | p-value |
| **Sex** |  |  | 0.002** |
|  | Female | 1 |  |
|  | Male | 1.17 (1.06, 1.29) |  |
| **Multiple birth** |  |  | <0.001*** |
|  | Singletons | 1 |  |
|  | Twins | 5.22 (4.52, 6.02) |  |
|  | Triplets | 21.5 (14.8, 31.2) |  |
|  | Quadruplets | 91.2 (31.6, 274) |  |
| **Birth rank** |  |  | 0.59 |
|  | 1 | 1 |  |
|  | 2 | 0.94 (0.84, 1.06) |  |
|  | 3 | 1.04 (0.89, 1.22) |  |
|  | ≥4 | 0.99 (0.76, 1.28) |  |
| **Marital status** |  |  | <0.001*** |
|  | Married<sup>#</sup> | 1 |  |
|  | Unmarried | 1.68 (1.50, 1.88) |  |
| **Nationality mother\*** |  |  | 0.09 |
|  | Switzerland | 1 |  |
|  | Southern Europe | 0.95 (0.77, 1.17) |  |
|  | EEA Europe | 1.08 (0.91, 1.27) |  |
|  | Non-EEA Europe | 1.15 (0.95, 1.39) |  |
|  | Other HIC | 1.23 (0.71, 2.14) |  |
|  | Other | 1.29 (1.07, 1.54) |  |
| **SEP index** |  |  | 0.04* |
|  | (Lowest) 1<sup>st</sup> quintile | 1.22 (1.02, 1.46) |  |
|  | 2<sup>nd</sup> quintile | 1.30 (1.09, 1.55) |  |
|  | 3<sup>rd</sup> quintile | 1.12 (0.94, 1.34) |  |
|  | 4<sup>th</sup> quintile | 1.21 (1.02, 1.43) |  |
|  | (Highest) 5<sup>th</sup> quintile | 1 |  |
| **Religion** |  |  | <0.001*** |
|  | Protestant | 1 |  |
|  | Catholic | 1.05 (0.91, 1.20) |  |
|  | Other religion | 1.31 (1.11, 1.55) |  |
|  | No religion | 0.76 (0.63, 0.91) |  |
| **Urban level** |  |  | 0.92 |
|  | Urban | 1 |  |
|  | Peri-urban | 1.01 (0.90, 1.12) |  |
|  | Rural | 0.98 (0.85, 1.13) |  |
| **Language region** |  |  | <0.001*** |
|  | German | 1 |  |
|  | French | 0.73 (0.65, 0.83) |  |
|  | Italian | 0.65 (0.48, 0.88) |  |

‡ Models additionally adjusted for maternal age, modelled by a quadratic spline with a knot at 30 years. # Includes registered partnership. \* We list countries in each category in Supplementary Table S1. Explanation of categories: EEA Europe, other countries in European Economic Area. Non-EEA Europe, European countries not in EEA. Other HIC, non-European high-income countries with low national infant mortality. Abbreviations: CI, confidence interval. RR, rate ratio. SEP, area-based socioeconomic position.

**Table S5:**
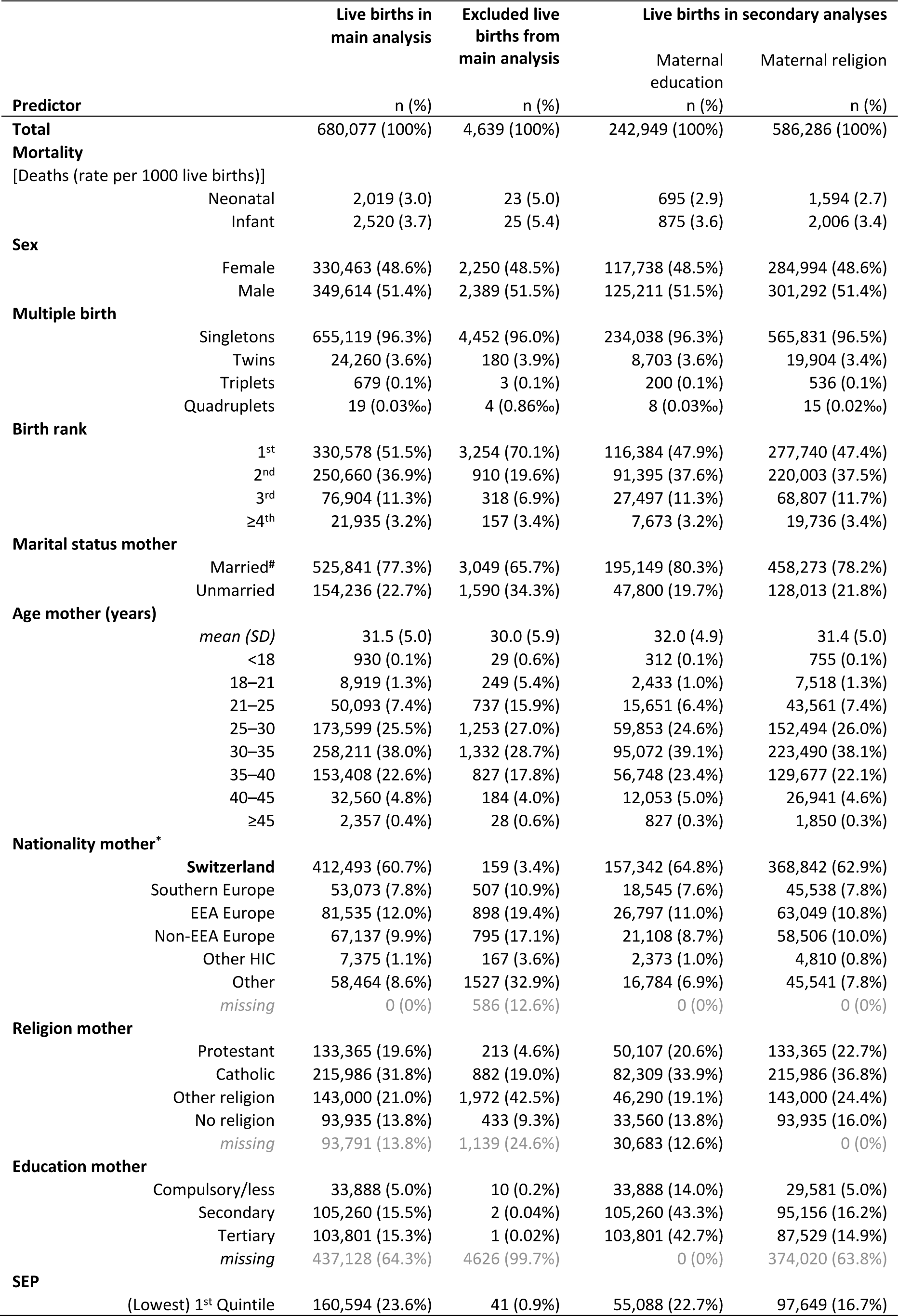
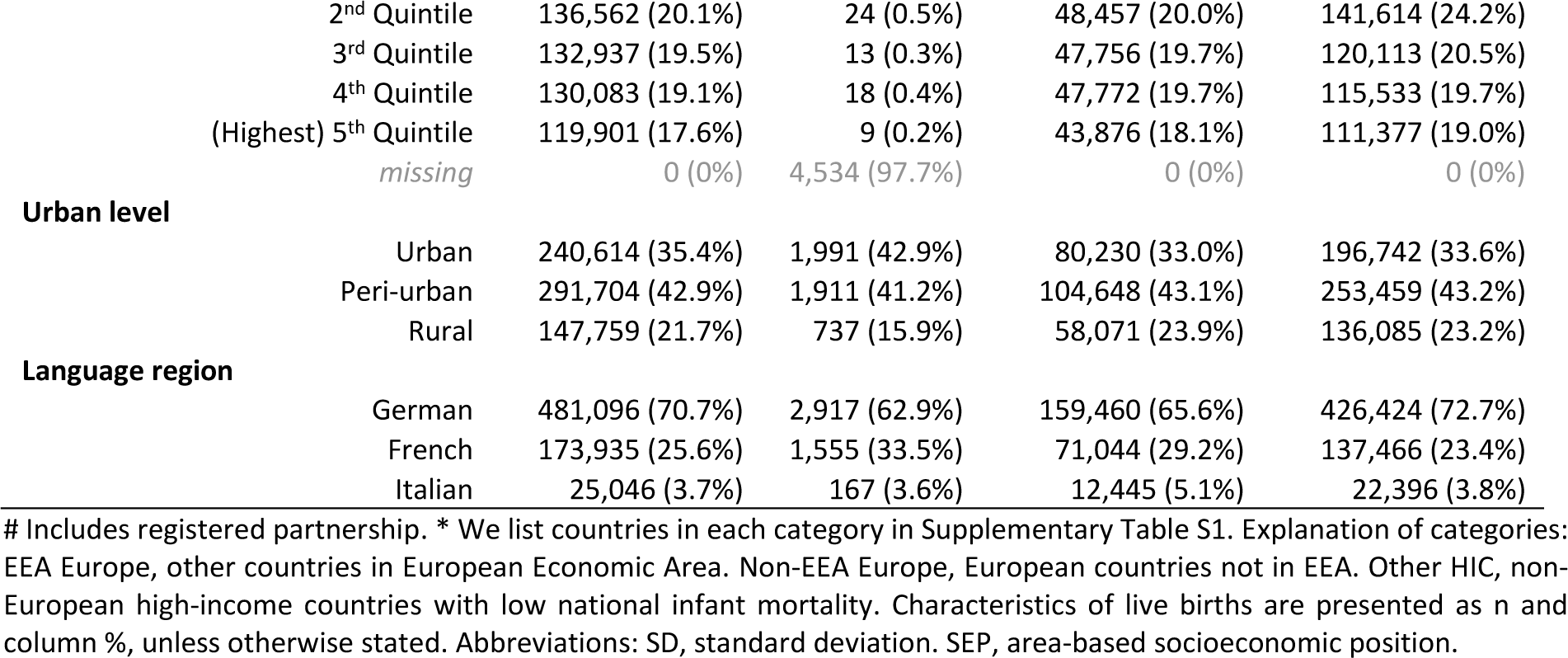
Population characteristics for live births included and excluded from main analysis and subsets of live births with available maternal education or available maternal religion (secondary analyses).

**Table S6.** Sensitivity analysis: Neonatal and infant mortality rate ratios (RR) in Switzerland 2011–2018, for all **singleton** live births (N = 655,119) based on multivariable Poisson models.

|  |  | Neonatal mortality |  | Infant mortality |  |
| --- | --- | --- | --- | --- | --- |
| Predictor |  | RR (95% CI) | p-value | RR (95% CI) | p-value |
| **Sex** |  |  | 0.038* |  | 0.002** |
|  | Female | 1 |  | 1 |  |
|  | Male | 1.11 (1.01, 1.22) |  | 1.14 (1.05, 1.25) |  |
| **Birth rank** |  |  | 0.08 |  | 0.009** |
|  | 1 | 1 |  | 1 |  |
|  | 2 | 0.91 (0.81, 1.02) |  | 0.97 (0.88, 1.07) |  |
|  | 3 | 1.06 (0.90, 1.25) |  | 1.11 (0.96, 1.28) |  |
|  | ≥4 | 1.18 (0.91, 1.52) |  | 1.40 (1.12, 1.74) |  |
| **Marital status** |  |  | <0.001*** |  | <0.001*** |
|  | Married# | 1 |  | 1 |  |
|  | Unmarried | 1.47 (1.31, 1.64) |  | 1.38 (1.25, 1.52) |  |
| **Age of mother‡** |  |  | <0.001*** |  | <0.001*** |
|  | 15 | 2.06 (1.09, 3.61) |  | 2.25 (1.35, 3.87) |  |
|  | 18 | 1.49 (1.03, 2.14) |  | 1.63 (1.21, 2.33) |  |
|  | 20 | 1.27 (0.98, 1.64) |  | 1.38 (1.13, 1.77) |  |
|  | 25 | 0.99 (0.92, 1.08) |  | 1.05 (0.99, 1.13) |  |
|  | 30 | 1 |  | 1 |  |
|  | 35 | 1.19 (1.08, 1.30) |  | 1.13 (1.04, 1.20) |  |
|  | 40 | 1.51 (1.34, 1.73) |  | 1.41 (1.27, 1.57) |  |
|  | 45 | 2.13 (1.57, 2.74) |  | 2.00 (1.57, 2.45) |  |
|  | 50 | 3.22 (1.84, 5.20) |  | 3.24 (1.98, 4.68) |  |
| **Nationality mother\*** |  |  | <0.001*** |  | <0.001*** |
|  | Switzerland | 1 |  | 1 |  |
|  | Southern Europe | 0.88 (0.71, 1.09) |  | 0.91 (0.75, 1.09) |  |
|  | EEA Europe | 1.07 (0.92, 1.25) |  | 1.06 (0.92, 1.22) |  |
|  | Non-EEA Europe | 1.46 (1.24, 1.72) |  | 1.36 (1.17, 1.57) |  |
|  | Other HIC | 1.36 (0.87, 2.13) |  | 1.45 (0.99, 2.13) |  |
|  | Other | 1.64 (1.41, 1.91) |  | 1.63 (1.43, 1.86) |  |
| **SEP index** |  |  | 0.25 |  | 0.02* |
|  | (Lowest) 1<sup>st</sup> quintile | 1.09 (0.92, 1.28) |  | 1.16 (1.00, 1.35) |  |
|  | 2<sup>nd</sup> quintile | 1.19 (1.01, 1.40) |  | 1.26 (1.09, 1.46) |  |
|  | 3<sup>rd</sup> quintile | 1.15 (0.98, 1.36) |  | 1.22 (1.05, 1.41) |  |
|  | 4<sup>th</sup> quintile | 1.07 (0.91, 1.26) |  | 1.11 (0.96, 1.29) |  |
|  | (Highest) 5<sup>th</sup> quintile | 1 |  | 1 |  |
| **Urban level** |  |  | 0.16 |  | 0.18 |
|  | Urban | 1 |  | 1 |  |
|  | Peri-urban | 1.09 (0.98, 1.22) |  | 1.06 (0.96, 1.17) |  |
|  | Rural | 0.98 (0.85, 1.13) |  | 0.95 (0.84, 1.08) |  |
| **Language region** |  |  | <0.001*** |  | <0.001*** |
|  | German | 1 |  | 1 |  |
|  | French | 0.72 (0.64, 0.81) |  | 0.81 (0.73, 0.90) |  |
|  | Italian | 0.66 (0.49, 0.89) |  | 0.74 (0.57, 0.95) |  |

**Table S7.**
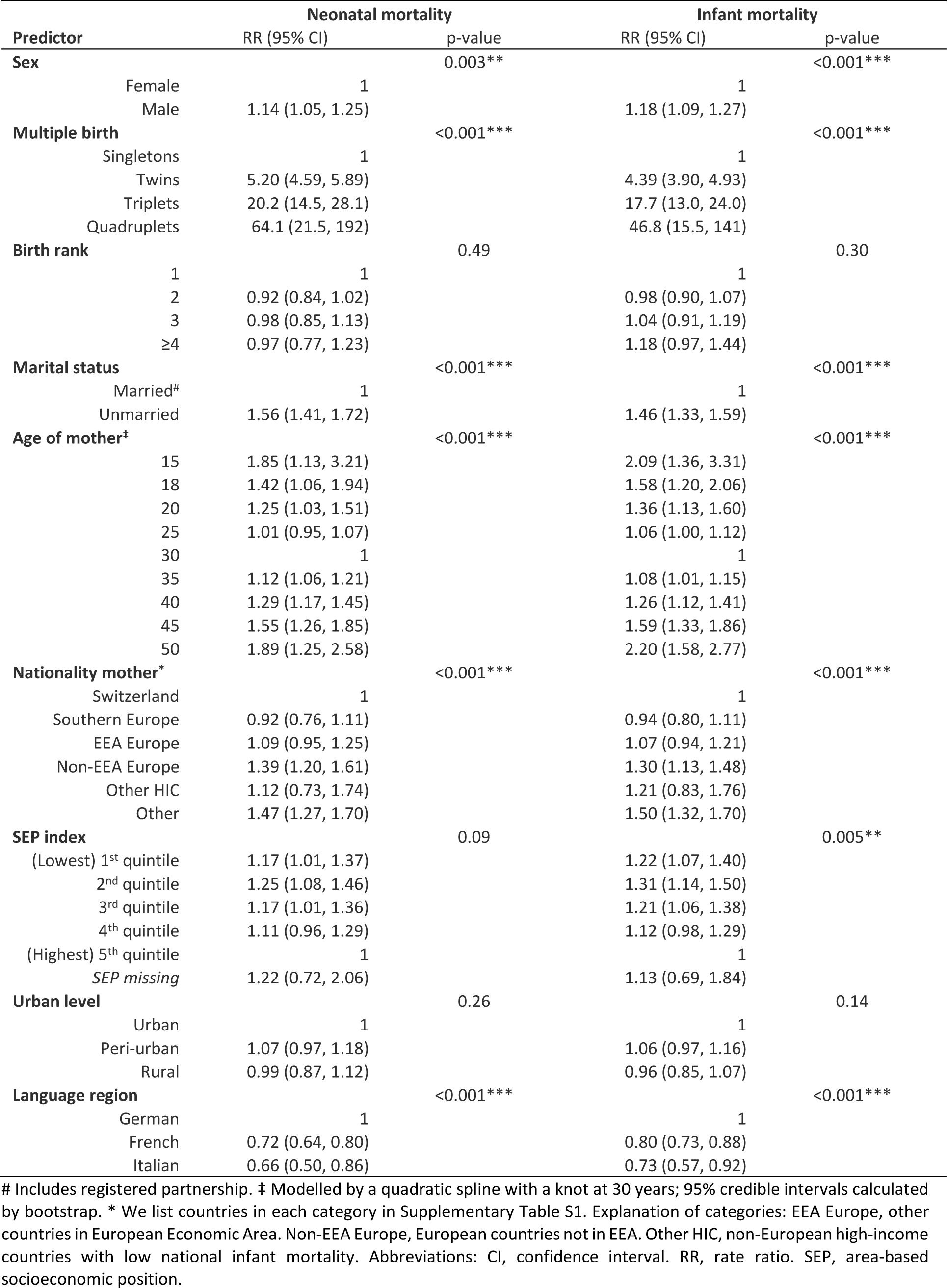
Sensitivity analysis: Neonatal and infant mortality rate ratios (RR) in Switzerland 2011–2018, for all live births including **6^th^ SEP category** for missing value (N = 684,130) based on multivariable Poisson models.

